# m^6^A-mRNA reader *YTHDF2* identified as a potential risk gene in autism with disproportionate megalencephaly

**DOI:** 10.1101/2022.12.21.22283275

**Authors:** Sierra S. Nishizaki, Nicholas K. Haghani, Gabriana N. La, Natasha Ann F. Mariano, José M. Uribe-Salazar, Gulhan Kaya, Melissa Regester, Derek Sayre Andrews, Christine Wu Nordahl, David G. Amaral, Megan Y. Dennis

**Author notes:** Corresponding author: Megan Y. Dennis, Ph.D. University of California, Davis School of Medicine, One Shields Avenue Genome Center, 4303 GBSF Davis, CA 95616.

## Abstract

Among autistic individuals, a subphenotype of disproportionate megalencephaly (ASD-DM) seen at three years of age is associated with co-occurring intellectual disability and poorer prognoses later in life. However, many of the genes contributing to ASD-DM have yet to be delineated. In this study, we identified additional ASD-DM candidate genes with the aim to better define the genetic etiology of this subphenotype of autism. We expanded the previously studied sample size of ASD-DM individuals ten-fold by including probands from the Autism Phenome Project and Simons Simplex Collection, totaling 766 autistic individuals meeting the criteria for megalencephaly or macrocephaly and revealing 153 candidate ASD-DM genes harboring *de novo* protein-impacting variants. Our findings include thirteen high confidence autism genes and seven genes previously associated with DM. Five impacted genes have previously been associated with both autism and DM, including *CHD8* and *PTEN*. By performing functional network analysis, we expanded to additional candidate genes, including one previously implicated in ASD-DM (*PIK3CA*) as well as 184 additional genes previously implicated in ASD or DM alone. Using zebrafish as a model, we performed CRISPR gene editing to generate knockout animals for seven of the genes and assessed head-size and induced-seizure-activity differences. From this analysis, we identified significant morphological changes in zebrafish loss-of-function of two genes, *ythdf2* and *ryr3*. While zebrafish knockouts model haploinsufficiency of assayed genes, we identified a *de novo* tandem duplication impacting *YTHDF2* in an ASD-DM proband. Testing zebrafish overexpressing *YTHDF2* showed increased head and brain size matching that of the proband. Single-cell transcriptomes of *YTHDF2* gain-of-function larvae point to reduced expression of Fragile-X-syndrome-associated FMRP-target genes globally and in the developing brain, providing insight into the mechanism underlying autistic phenotypes. We additionally discovered a variant impacting a different m^6^A-methylation reader, *YTHDC1*, in our ASD-DM cohort. Though we highlight only two cases to date, our study provides support for the m^6^A-RNA methylation pathway as potentially contributing to this severe form of autism.

**Lay Summary:** Autism (ASD) has become increasingly prevalent in children in recent years due to increasing awareness and improving diagnosis. While we know ASD is associated with hundreds of genes, there is much to learn about what genes are involved and how they contribute to its diverse presentation of traits and behaviors. By focusing on autistic individuals exhibiting enlarged brains (disproportionate megalencephaly), we identified new candidate genes possibly contributing to ASD and brain size. A autistic-patient-identified duplication of one gene in particular, *YTHDF2*, implicates a novel pathway related to RNA modifications with ASD and human brain size for the first time.

## INTRODUCTION

Autism is a group of neurodevelopmental traits characterized by difficulties with communication, social interactions, and behavioral challenges, prevalent in 1 out of 36 children in the United States [1]. Autism is highly heritable, with 50–90% of cases estimated to be driven by genetics alone [2–4]. Autism is also highly heterogeneous with large-scale whole exome sequencing (WES) of >63,000 autistic probands identifying 125 high confidence autism genes, with the predicted number of genes left to be discovered exceeding 1,000 [5–7]. In particular, leveraging genomic data from autism families—including parents and unaffected siblings—in the Simons Simplex Collection (SSC) [8] has identified coding *de novo* variants estimated to contribute to 30% of diagnoses [9–11]. More recently, whole genome sequencing (WGS) of SSC *de novo* noncoding mutations implicates risk in an additional 4.3% of autism cases [12,13]. Despite the combined efforts to sequence tens of thousands of genomes, known genes still only account for 5-20% of cases, and further work is required to fully elucidate genes and pathways contributing to autism etiology [5,6,14–22]. Combining *de novo* variation with autism sub-phenotyping has been used to address the heterogeneity of autism and identify susceptibility loci for comorbid phenotypes in an acute way [23].

Brain enlargement that is disproportionate to height, known as disproportionate megalencephaly (DM), is enriched in autistic probands with 15% of autistic boys falling under the DM subphenotype (ASD-DM) compared to 6% in typically developing (TD) boys [24]. This comorbidity is associated with more severe cognitive phenotypes, including lower IQ and language use, as well as higher rates of language regression [25–27]. This robust enrichment and distinct presentation support DM as a sub-phenotype of ASD, likely due to a shared genetic etiology between autism and DM. While a handful of genes have been associated with DM—including known autism genes impacting cell cycle and proliferation during embryonic development (*e.g.*, *CHD8* and *PTEN*)—mutations of known candidate genes make up only 3% of megalencephaly in autism probands. This leaves the genetic etiology of a majority of ASD-DM cases undiscovered [28–31]. A study using WES from 46 autistic families with macrocephaly (ASD-M)— defined as >2 standard deviations above the mean in head circumference for TD sex and age-matched children—successfully identified mutations in one novel and several known autism candidate genes [32], demonstrating the power of sub-phenotyping ASD-DM leading to genetic discoveries even for reduced sample size.

Zebrafish (*Danio rerio*) are an attractive model for studying neurodevelopmental traits given their rapid development, large number of progeny, transparent bodies, and that ∼70% of gene orthologs are shared with humans [33–35]. Previous studies of known ASD-DM genes recapitulate macrocephaly and DM phenotypes in zebrafish knockdown and knockout experiments for *CHD8* and *KMT2E*, respectively [36,37]. This method of knocking down candidate ASD-M genes has also been used systematically to identify the contributing gene in the chromosome 16p11.2 locus in zebrafish [38]. Further, novel technologies such as the VAST BioImaging System allow for the rapid characterization of zebrafish knockout models through the generation of high resolution standardized images [39]. We recently demonstrated the utility of this approach by examining the knockdown of two genes associated with autism and microcephaly, *SLC7A5* and *SYNGAP1*, by assessing head-size phenotypes in CRISPR-generated zebrafish knockout line embryos at three and five days post fertilization (dpf) [40].

In this study, we leveraged high-coverage WGS data from two cohorts, 11 ASD-DM probands from the UC Davis MIND Institute Autism Phenome Project (APP) specifically identified using magnetic resonance imaging (MRI) data at around three years of age, and 755 ASD-M probands with head circumference data available from the SSC cohort. Together, this represents a >10-fold increase in probands compared to the previous largest study of increased head circumference associated genes in ASD [32]. Using this sub-phenotype-to-genotype analytic strategy, we identified candidate ASD-DM and ASD-M genes harboring *de novo* likely gene-disrupting (LGD) variants, including *CHD8* and *PTEN*, and subsequently functionally validated a subset of candidate genes using zebrafish. From this, we narrowed in on two genes impacting larval head size. Overexpression of *YTHDF2* results in macrocephaly in zebrafish following embryonic microinjection of mRNA, recapitulating the phenotype seen in the ASD-DM proband harboring a partial tandem duplication of the gene. Together our sub-phenotyping approach provides a powerful strategy to identify novel ASD-DM candidate genes and validate their role in brain development using a zebrafish model system.

## METHODS

### Megalencephaly and Macrocephaly Phenotypes

APP probands and determinations of megalencephaly were previously determined as part of the APP study [24]. Acquisition of MRI data for megalencephaly measurements were made during natural nocturnal sleep for children at study enrollment (time point 1), between the ages of 2 and 3.5 [41]. Blood collected during this time point was sequenced using the to 30X coverage WGS through a collaboration with MSSNG [42,43]. Raw data including FASTQ and VCF files can be accessed through the MSSNG access agreement: https://research.mss.ng.

Macrocephaly cases from the SSC were defined using a permissive cutoff of a head circumference >1.5 standard deviations (90%) above the mean of age matched controls [44]. Age matched TD head circumference data [45] and height data [46] from ages 4-17 were derived from publicly-available standards. For males in the SSC cohort 551/1601 (34%) met the criteria for macrocephaly, for females 108/245 (45%) met this criteria. We identified three types of macrocephaly: (1) somatic overgrowth (SO) with head circumference and height percentiles >90%; (2) disproportionate macrocephaly (DM) with height percentiles over head circumference percentile < 0.7; and (3) relative macrocephaly (RM) with height percentiles over head circumference percentile > 0.7.

### Variant Annotation

Whole-genome sequencing, read mapping, and variant identification was performed for APP families as part of the MSSNG consortium [42,43]. *De novo* variants were identified in APP probands as those unique to the proband and absent from either parent via string matching (grep -Fvxf). We considered LGD variants as those predicted to lead a frameshift, nonsense, or splice site mutation. Rare variants were identified using dbSNP as those with a minor allele frequency (MAF) < 0.2% in all five 1000 Genomes ancestry-based populations [47,48]. The presence of all rare and *de novo* variants identified in the APP cohort were validated by visual inspection of sequencing data via the Integrated Genomics Viewer (IGV) [49].

### Network and Ontology Analyses

Network analysis of known ASD-DM genes and candidate ASD-DM genes from this study were completed using the STRING database and visualized via Cytoscape [50,51]. Gene ontology (GO) analysis was completed for known ASD-DM genes and candidate ASD-DM genes as seed genes along with their top ten gene interactors determined using the STRING database, similar to previous studies [31]. Similar STRING database Molecular Function GO terminology were pooled. GO was completed using Database for Annotations, Visualization, and Integrated Discovery (DAVID) software [52]. Human genes were used as background for GO analyses.

### Zebrafish CRISPR and mRNA models

Guide RNAs (gRNAs) were selected as having a CRISPRScan score of 35 or higher (Table S1) [53]. crRNA were synthesized by Integrated DNA Technologies. Injection mixes of ribonucleic protein (RNP) consisting of four pooled gRNA (annealed crRNA and tracrRNA) and SpCas9 Nuclease (New England Biolabs, M0386M) in order to achieve 90% knockdown efficiency [54], and were prepared as previously described [40]. Pooled gRNA (4 μM total concentration) were microinjected into single cell NHGRI-1 or transgenic zebrafish embryos to a volume of 0.5 nL/cell as previously described using a Pneumatic MPPI-2 Pressure Injector [55]. Scrambled injection RNP mix contained a single gRNA designed to have no target in the zebrafish genome. gRNA efficiencies were tested post-injection using pooled genomic extractions of four embryos and PCR amplification of targeted loci followed by 7.5% polyacrylamide gel visualization. These same amplicons were also subject to Illumina sequencing and the total alleles identified/quantified using the CrispRvariants R package (Figure S1, Table S1) [56].

Human mRNA was generated using cDNA plasmids (Horizon *YTHDF2*, MHS6278-202827242; *GMEB1*, MHS6278-202827172) [57] and prepared via the *in vitro* transcription kit mMESSAGE mMACHINE™ SP6 Transcription Kit (Thermo Fisher Scientific, AM1340). Mixes of 100 ng/μL mRNA and 0.05% phenol red were prepared, as previously described, and injected in single-cell zebrafish embryos at a volume of 0.5 nL/cell [58].

### Zebrafish Morphometric Measurements

Dorsal and ventral images of 3 days post fertilization (dpf) embryos were obtained using the Union Biometrica VAST Bioimaging System with LP Sampler via the built-in camera and manufacturer settings [39]. Zebrafish features were identified and quantified from VAST using FishInspector software 2.0 [59]. FishInspector images were assessed for total area (contourDV_regionpropsArea), embryo length (contourDV_regionpropsLengthOfCentralLine), distance between the center of the eyes (YdistanceCenter_eye1DV_eye2DV), and telencephalon distance (YdistanceEdge_eye1DV_eye2DV). Statistical analysis was performed in R using the ggsignif package using the Wilcoxon test option [57,60].

Fluorescent images were acquired using the Andor Dragonfly High Speed Confocal Platform with the iXon Ultra camera. Human *YTHDF2* mRNA wasd microinjected Tg(HuC-eGFP) strain zebrafish, which harbor a green-fluorescent protein (GFP) fluorescent pan-neuronal marker, were bathed in 0.003% 1-phenyl-2-thiourea (PTU) in 10% Hank’s saline between 20–24 pf for 24 hr (Fisher Scientific, 5001443999). At 3 dpf zebrafish embryos were embedded in 1% low melt agarose (Thermo Fisher Scientific, BP160-100) and imaged using a GFP filter.

### Zebrafish Seizure Analysis

Zebrafish movement was recorded at 30 frames per second for 1 hr with no stimulation using ViewPoint’s ZebraBox Technology per manufacturer recommendations [61]. Embryos were maintained at 37°C using a polystat water heating system. Movements were detected over one second intervals. Distance and time moved were analyzed using a custom R script and high speed events were quantified using a previously published MATLAB script [62].

### Zebrafish RNA Extraction and RT-qPCR

Whole zebrafish larvae were collected at 3 dpf and stored in 50 μL RNALater at -80°C until RNA extraction. Three biological replicate samples were prepared for both *ythdf2* KO and scrambled control larvae, each containing 15 larvae, for pooled RNA extraction using an RNeasy Plus Mini kit (Qiagen). Briefly, larvae were resuspended in 350 μL of the buffer RLT and vortexed until homogenized. Instructions from the RNeasy Plus Mini kit were followed for DNA Removal using the gDNA Eliminator column. Samples were quantified using a Qubit BR kit and normalized to 4 ng/μL for RT-qPCR following the instructions from the NEB Luna kit.

### Single-cell transcriptomics

Transcriptional differences across *YTHDF2* zebrafish models were assessed using single-cell (sc)RNA-seq. Cells were prepared from *ythdf2* knockout and SpCas-scrambled gRNA controls as well as *YTHDF2*-injected and eGFP-mRNA-injected controls. At 3 dpf, larval heads from each group were dissected after euthanasia in cold tricaine (0.025%), pooling 15 heads together per sample with three samples per group. Groups with low initial counts (*ythdf2* knockout and eGFP-mRNA) were repeated with an additional three samples. Cells from each sample were washed with 1 ml of cold 1x PBS twice and immediately incubated at 28°C in a mix of 480 μl of Trypsin-EDTA (0.25%) and 20 μl of Collagenase P (100 mg/ml) for a total of 15 min with gentle pipetting every 5 min to induce dissociation. To stop dissociation, 800 μl of cold DMEM with 10% FBS was mixed with each sample and immediately centrifuged at 4°C for 5 min at 700*g*. The supernatant was carefully removed from the cell pellet and cells were washed in cold 1x PBS and centrifuged at 4°C for 5 min at 700*g*, followed by another wash of cold DMEM with 10% FBS. Cells were then filtered into Eppendorf tubes using a P1000 pipette and a Flowmi 40 µm cell strainer (Sigma Aldrich, St. Louis, MO). 10 μl of sample was then mixed with 10 μl of trypan blue solution and counted using a Countess II (Thermo Fisher, Waltham, MA) to record cell viability. All samples processed were confirmed to show viability above 70%.

Cell fixation and library preparation were performed following the sci-RNA-seq3 protocol [63] using DSP/methanol. After the combinatorial indexing and PCR amplification steps, all wells were pooled together to ensure sufficient library yield before purification. The pooled libraries were then purified using AMPure XP beads to remove any remaining small fragments and primers. The quality and concentration of the libraries were assessed using a Bioanalyzer (Agilent Technologies, Santa Clara, CA) to ensure they met the required size distribution and concentration thresholds. Final libraries were size-selected using the Pippin HT system (Sage Science, Beverly, MA). The target range was set to 400-500 bp, with the smear cut between approximately 300-600 bp to ensure that only fragments within this desired range were included. The libraries were sequenced with paired-end read length of 150 bp using the Illumina NovaSeq 6000 platform.

FASTQ files were processed according to the sci-RNA-seq3 bioinformatic pipeline (https://github.com/JunyueC/sci-RNA-seq3_pipeline) and a comprehensive zebrafish transcriptome [64] was used to generate cell-by-gene matrices per sample. These matrices were processed into Seurat objects using *Seurat* v5.0.3 [65]. Cells with mitochondrial or ribosomal percentages above 5%, feature counts below 200 or over two standard deviations from the mean, and predicted doublets according to *DoubletFinder* [66] were removed from subsequent analyses. After quality-control filtering, an average of 1,126 cells per sample (4,785 cells per group) were obtained and normalized with the 5,000 most variable genes while regressing for ribosomal and mitochondrial percentages using *SCTransform*.

Samples were integrated using a reciprocal PCA reduction [65] and nearest-neighbor graphs were made using the first 30 principal components with the *FindNeighbors* function for subsequent clustering. Hierarchical clustering was initially performed using the Euclidean distance between all cells from principal component embeddings with the tree cut at k = 10. Broad marker genes were assigned using the *PrepSCTFindMarkers* and *FindAllMarkers* functions using the wilcox test option (parameters: *logfc*.*threshold*= 0.1, *min.pct*= 0.1, *return.thresh*= 0.01, *only.pos*= TRUE). Brain cells from a single broad cluster were isolated and hierarchical clustering was similarly repeated with the tree cut at k = 18. Cell clusters were defined using defined marker genes cross-referenced with larval zebrafish brain atlases [67,68] and the Zebrafish Information Network (ZFIN) [69].

For the differential gene expression analysis, cells from the *ythdf2* knockout and *YTHDF2*-injected groups were randomly sampled with respect to our original cluster distribution to match control cell counts (*downsampleSeurat*). Differentially-expressed genes (DEGs) were identified across all and a subset of brain cells, respectively, using the *FindMarkers* function with the wilcox test option (parameters: *logfc*.*threshold*= 0.1, *min.pct*= 0.01, *only.pos*= FALSE). A list of 842 high-confidence Fragile X Syndrome (FXS) protein (FMRP) targets [70] were converted to zebrafish orthologs using the g:Orth search from g:Profiler [71] to identify FMRP-target DEGs (adjusted p-value < 0.05) and assess enrichment using a Benjamini Hochberg (BH)-adjusted Fisher’s exact test. All other FMRP-target genes expressed across both *ythdf2* knockout and *YTHDF2* mRNA conditions in at least 0.01 percent of cells were selected for subsequent analysis (n=675). *scCustomize* [72] and *dittoSeq* [73] were used for figure creation. *Nebulosa* [74] was used to visualize joint expression from multiple FMRP-DEGs using a kernel gene-weighted density estimation.

## RESULTS

### ASD-DM Candidate Gene Discovery

ASD-DM individuals were recruited through the UC Davis APP—a longitudinal study focused on the identification of ASD-subphenotypes [24,75,76]. Using MRI data from the study entry time point (2–3½ years of age) [75], we selected 11 individuals in the APP cohort that met the criteria for ASD-DM, defined as a cerebral volume to height ratio >1.5 standard deviations above the mean compared to TD age-matched controls. Through a collaboration with MSSNG [17,43], WGS and variant identification/annotation was performed for the autistic probands and a subset of family members, for which we also had blood specimens, including six trios and five non-trio probands yielding over 200 thousand variants. From this, we identified two exonic, *de novo*, LGD variants from trio families, including one single-nucleotide variant (SNV) splice-site variant impacting *RYR3* and one 109-kbp duplication of *YTHDF2* and *GMEB1*. From the five individuals with no parental data, we identified a proband harboring a chromosome 1q21.1 microduplication, a copy-number variant (CNV) previously associated with ASD-DM [77], and a single proband with variants in *CHD8* and *KMT2E* [78]. An additional ten variants were found in non-trio proband data to be exonic, LGD, and rare (not previously recorded in dbSNP) [79]. Of these, three impacted genes have SFARI scores of 3S or above (*KMT2E*, *RPS6KA5*, and *TTN*), an additional three genes have known neuronal functions (*DMBT1*, *IARS2*, *FGF12*), and one gene was found recurrently carrying variants in two probands (*SPANXN4*) [80].

SSC consists of trios and quads of simplex autism families with accompanying genetic and phenotypic information. Due to the lack of MRI data for SSC participants, we used ASD-M as a proxy for ASD-DM. SSC head circumference and age data was used to determine ASD-M status (head circumference >1.5 standard deviations above the mean for typically developing sex and age-matched children) for 755 of 1,846 SSC probands (40%) [8] (Table S2, Figure S2). Considering only SNVs and indels, ASD-M *de novo* LGD variants were identified from published results [12], overlapping a total of 150 genes (Figure 1, Table S3). Of note, five genes were found recurrently mutated in ASD-M, including *GALNT18*, *KDM6B*, *LTN1*, *RERE*, and *WDFY3*, as well as *CHD8*, which was disrupted in three probands.

**Figure 1.**
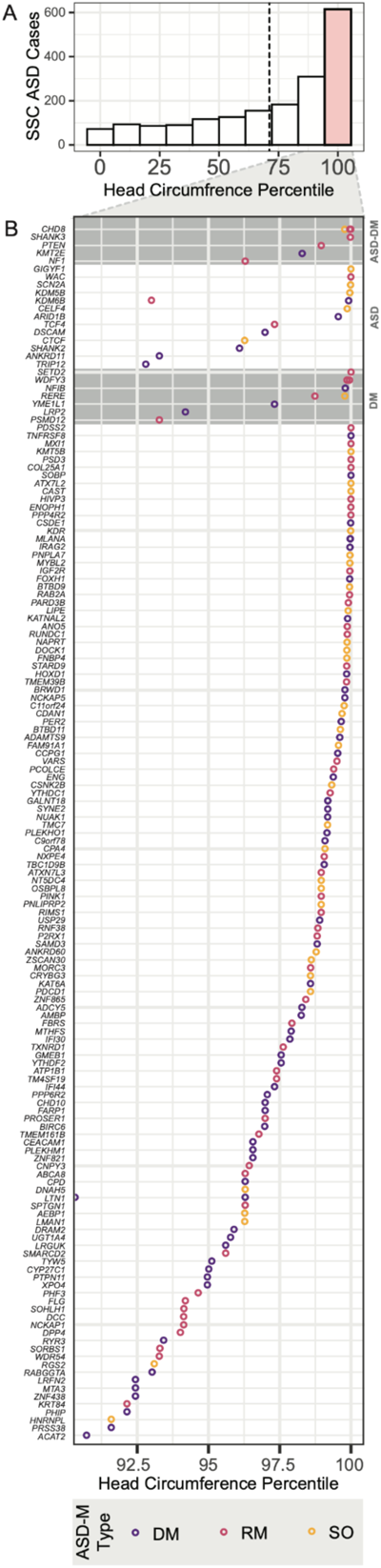
Macrocephaly level of candidate ASD-DM and ASD-M genes. **(A)** A histogram representing the number of SSC probands v. head circumference percentiles shows a skew towards larger head-sizes compared to age and sex-matched typically developing children. The red bar designates those meeting the criteria for macrocephaly. The dashed line represents the distribution mean. **(B)** ASD-DM and ASD-M genes listed by their identified proband’s head circumference percentiles show genes previously associated with ASD-DM (first gray quadrant) are more likely to be associated with a higher head circumference percentiles than genes previously associated with autism (second white quadrant) and DM (third gray quadrant) alone. Color represents the macrocephaly type. DM, disproportionate macrocephaly; RM, relative macrocephaly; SO, somatic overgrowth.

In total, we identified 153 genes containing a LGD variant across the APP ASD-DM and SSC ASD-M datasets. Rates of harboring a LGD *de novo* variant in ASD-DM probands were in line with previous predictions (19.4%) and nominally enriched compared to TD SSC siblings (16.5%), though not statistically significant (chi-squared p-value = 0.1) [9,32]. Over a third of identified candidate genes (53/153) had a pLI score of > 0.9, suggesting intolerance to variation [81] (Table S3). Examining *de novo* missense variants, which were previously shown to exhibit overall enrichments in affected probands versus unaffected siblings [82,83], we did not observe an enrichment in our LGD candidate genes in ASD-M probands compared to TD siblings of ASD-M probands, ASD-without-macrocephaly (ASD-N) probands, and siblings of ASD-N probands (Student’s T-test, p-values = 0.16, 0.29, 0.76).

### ASD-DM Candidate Gene Discovery Using Network Analysis

Identifying shared patterns of molecular functions and ontologies of impacted ASD-DM genes may point to additional gene candidates. Due to the highly heterogeneous nature of ASD, this type of analysis expands our ability to identify disrupted biological mechanisms and spatio-temporal expression patterns implicated in autism [31]. Here, we used as seeds 166 previously-known and identified-in-this-study ASD-DM genes to identify active interactions using the STRING database (Figure 2) [32,50]. This analysis uncovered ontology groups enriched in our dataset previously reported for ASD, including proteins involved in histone modification and chromatin organization, transcription factors, cell signaling (e.g., SMAD and E-box binding), functions key to neuronal activity (e.g., sodium and calcium ion transport and glutamate receptor binding), cell adhesion and cytoskeletal proteins, and mRNA binding (Table S4) [84–87]. Out of our original ASD-DM candidate seed genes 41.5% (69/166) fall under one of these ontologies.

We next used the Database for Annotations, Visualization, and Integrated Discovery (DAVID) to identify unique ontologies enriched in the 166 known and candidate ASD-DM genes compared to ontologies enriched in SSC ASD-N proband LGD genes, versus genes previously associated with DM [52,88]. While there are many commonalities between the subphenotype and ASD-N, including chromatin remodeler, ASD-DM is uniquely enriched for terms such as intellectual disability, synapse assembly and long-term synaptic depression, histone methyltransferase activity, cytoskeletal structure (spectrin repeats) (Table S4). Notably, unlike ASD-N (*GRB10*, *PPP2R5D*, *RICTOR*, and *TSC1*), there was no enrichment in LGD variants impacting genes related to the MTOR pathway for ASD-DM (*NF1* and *PTEN*) [89].

We next sought to identify putative additional candidate ASD-DM genes by expanding our network to include the top ten interactors for each ASD-DM seed gene (Table S5). This list of top ten interactors included genes defined by STRING as having known protein interactions, shared homology, and co-expression patterns [50]. Of the ASD-DM candidate gene interactors, 28% (518/1826) fall under one of the ontologies found in our ASD-DM network. Interestingly, one of these genes has previously been associated with both autism and DM individually, *PIK3CA* (Table S6). *PIK3CA* functions as a catalytic subunit of the mTOR pathway and has previously been found to be associated with developmental delay and DM, including one individual diagnosed with autism [90].

In this ASD-DM interactor set, twenty genes are high-confidence autism genes not previously associated with DM (*ANK2*, *ASXL3*, *CTNNB1*, *CUL3*, *DLG4*, *DYRK1A*, *GNAI1*, *GRIN2B*, *KCNMA1, KMT2A*, *NCOA1*, *NIPBL*, *NRXN1*, *PHF12*, *POGZ*, *PPP1R9B*, *SIN3A, SMARCC2*, *TBL1XR1*, *UBR1*). Eighteen additional genes from this interactor set have been implicated in DM and implicated as SFARI putative autism candidate genes (*ANK3*, *CHD2*, *CHD3*, *FRMPD4*, *HCFC1*, *HDAC4*, *HRAS*, HUWE1, *PAK1*, *PIK3R2*, *RAC1*, *SETD1A*, *SLC25A1*, *SMAD4*, *TBL1X*, *TRIO*, *USP7*, and *USP9X*), and 184 more have been implicated in DM or have a SFARI score. Especially promising are the 21 genes that contain missense variants in the SSC ASD-M probands, but not in ASD-N probands or their TD siblings, including *ABI2*, *ANK3*, *SRC*, *SRCAP*, *ATP12A*, *BAIAP2*, *CHD13*, *CH815*, *FGG*, *JUP*, *KDM2A*, *KIF20A*, *MAPK8*, *PDGFRB*, *RING1*, *SCN4A*, *SHANK1*, *SMC3*, *TCF3*, *WDR5*, *ZC3H3*. Together, network analysis and ontology point to these genes as promising ASD-DM candidate genes going forward.

**Figure 2.**
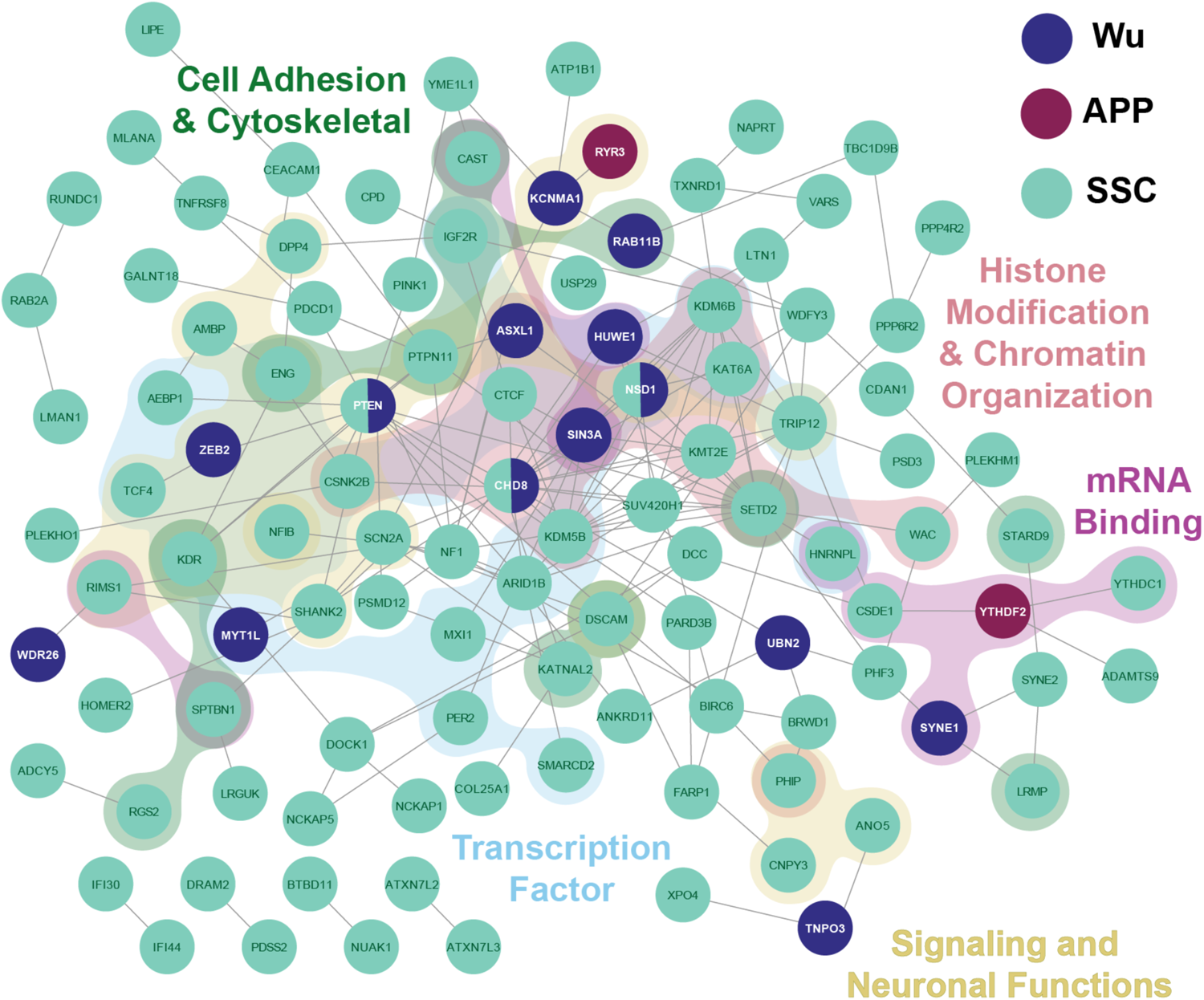
Network analysis and gene ontology of ASD-DM candidate genes. ASD-DM candidate genes from SSC (teal), APP (purple), and Wu (navy) probands are connected in a network via active interactions as determined by STRING [50]. Background colors represent shared GO molecular functions. Disconnected gene nodes are not included.

### Validation of Candidate Genes in a Zebrafish Model System

In order to determine if our candidate ASD-DM genes contribute to a head-size phenotype, we tested seven by CRISPR knockout in zebrafish (Figures S1 & S3). We focused our analysis on the previously unstudied candidates from the APP cohort, including the three genes impacted by *de novo* variants, *RYR3*, *GMEB1*, and *YTHDF2*. It is known that ∼20% of zebrafish genes conserved with humans are also duplicated; we therefore prioritized candidate ASD-DM genes with only a single ortholog in zebrafish (*IARS2* and *RPS6KA*) [91]. Additionally, we included two genes from the SSC ASD-M cohort, *CHD8*, for which CRISPR F_0_ mosaic knockouts have previously been shown to lead to increased interorbital distance in zebrafish embryos [36], and *FAM91A1*, a gene previously not implicated in ASD. Finally, as controls, we included three genes found in SSC ASD-N probands for which we do not expect to see a head-size phenotype—*HEPACAM2*, *PAX5*, and *SCP2A*.

We generated CRISPR knockout F_0_ embryos (or “crispants”) by microinjection of four gRNAs targeting exonic regions of each candidate gene. This approach has been shown to result in near complete mosaic knockout of genes with little off-target effects [32,40,92]. We then quantified morphometric features in 3 dpf larval crispants, including embryo area, body length, and distance between the center of the eyes (as a proxy for head size). From this analysis, we identified two genes (*ryr3* and *ythdf2*) showing significant head-size differences in zebrafish knockout embryos compared to negative scrambled injection controls (Wilcoxon T-test p-values < 0.01) (Figure 3). In both cases, crispant embryos exhibited reduced head size consistent with microcephaly, with *ryr3* showing a mean reduction of 3.3% and *ythdf2* showing a mean reduction of 3.8%. *RYR3* encodes a ryanodine receptor responsible for calcium transport in muscle and brain tissue, and *YTHDF2* encodes for part of the m^6^A-mRNA degradation complex [93,94]. Though *ythdf2* also displayed a reduction of body area and length, this size difference did not appear to be due to overall developmental delay, measured by head-trunk angle (Figure S4). This was in contrast to a previous study of a morpholino *ythdf2* zebrafish knockout, which noted an overall delay in development as well as potential compensation of m6A-mRNA clearance activity [95]. Only genes harboring known *de novo* variants from the ASD-DM APP cohort exhibited head-size phenotypes, though we did not observe any morphometric differences of our F_0_ mosaic *chd8* crispant, counter to published results in stable lines and using morpholinos [35,36,96].

As increased prevalence of seizures is highly enriched in autism [97], we assessed our knockout embryos for increased susceptibility to drug-induced seizures [98] by treating them with GABA antagonist pentylenetetrazole (PTZ; 5 mM) at 5 dpf [40,99]. High-speed events, corresponding to seizure-like movements, were recorded using motion tracking [61] and quantified [62] (Figure 3). This analysis revealed three crispant models associated with increased high speed events versus scrambled controls, *fam91a1*, *gmeb1*, and *hepacam2*. Of note, *HEPACAM2,* an immunoglobulin gene responsible for cell adhesion, showed the highest fold increase in high speed events compared to controls in our zebrafish assay. This gene falls within the human chromosome 7q21.3 microdeletion syndrome region associated with myoclonic seizures in humans [32,100]. Mutations in *FAM91A1*, responsible for Golgi protein trafficking, have been linked to dysregulated electrophysiological brain activity [101,102], while *GMEB1*, a caspase activation and apoptosis inhibitor, has no previous connections with seizures.

**Figure 3.**
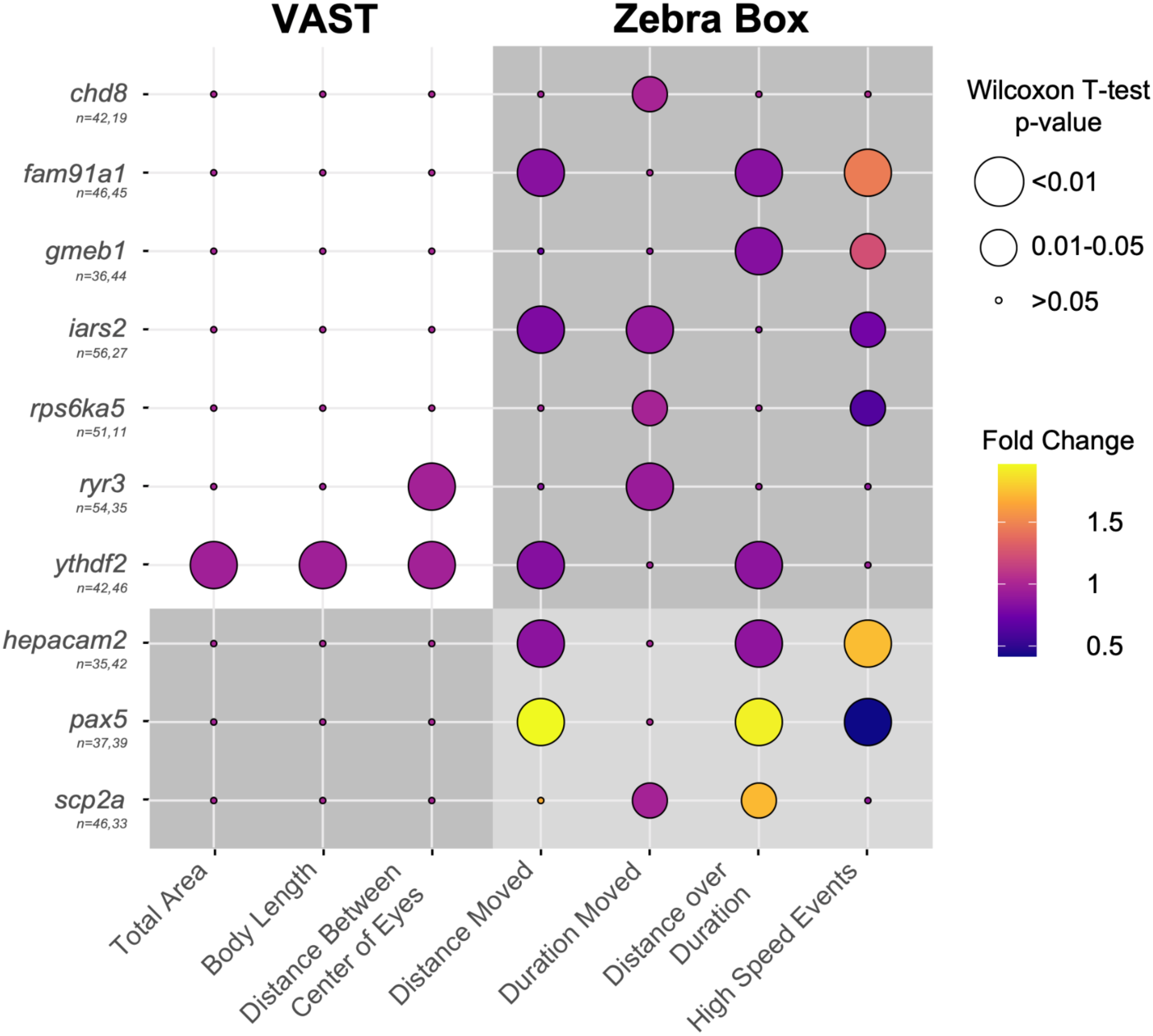
Zebrafish CRISPR knockout embryos for the rapid validation of ASD-DM genes. Phenotyping knockout zebrafish morphometric measurements using the VAST BioImaging System, we identified *ryr3* and *ythdf2* as having reduced head size compared to a negative scrambled control. Seizure-like activity in the presence of PTZ was assessed using ZebraBox video motion tracking. Circle size represents the inverse Wilcoxon T-test p-value. Circle color represents the fold change compared to a negative scrambled control.

### *YTHDF2* in ASD-DM

Based on the findings that knockout crispants targeting *ythdf2* exhibited microcephaly and *gmeb1* displayed increased seizure activity, we sought to better characterize the *de novo* 109-kbp duplication identified in an APP ASD-DM proband. To begin, we validated the duplication existed in the proband but not parents using sequence read depth (QuicK-mer2) [49,103] (Figure 4A). Split reads falling at the identified breakpoints indicated that the duplication, harboring the entire *GMEB1* gene and the first five of six exons of *YTHDF2*, inserted in tandem at the 3’ untranslated region (UTR) of the noncoding divergent transcript of *TAF12*, directly upstream of *GMEB1* (Figure 4B and C). Using available microarray data produced from mRNA derived from whole venous blood, we found that both *GMEB1* and *YTHDF2* exhibited increased expression > 3 standard deviations from the mean in the APP proband harboring the duplication compared to other APP participants [104].

*GMEB1* is an auxiliary factor in parvovirus replication known to inhibit apoptosis in neurons and previously associated with schizophrenia [105,106], and *YTHDF2* is a member of the m^6^A-containing mRNA degradation complex known to be downregulated in neuronal fate determination [94], making both of these attractive potential ASD-DM candidate genes. Based on their known functions, duplication of either gene could plausibly result in neurodevelopmental effects. Therefore, we modeled increased expression of *YTHDF2* and *GMEB1* by microinjecting human *in vitro* transcribed mRNA into single-cell stage zebrafish embryos. We then assessed morphometric features of ‘overexpression’ zebrafish embryos at 3 dpf (Figure 4D). For *YTHDF2*, we observed increased telencephalon length compared to a dye injected negative control consistent with increased head size but no significant differences to body area or length, matching the proband phenotype. We did not observe any difference in head size for the *GMEB1*-injected larvae (Figure 4D). To verify the increased head size was a result of an enlarged brain, we repeated the experiment in the zebrafish transgenic line HuC-GFP [107], which harbors a green-fluorescent protein (GFP) pan-neuronal marker (Figure 4E and F). These embryos displayed brain size differences, with knockout embryos showing significantly reduced midbrain, and mRNA injected ‘overexpression’ embryos exhibiting significantly increased midbrain and forebrain compared to injection controls. Together, the knockout and mRNA injected zebrafish provide evidence that increased dosage of *YTHDF2* is associated with DM while its loss leads to microcephaly.

Exploring the *YTHDF2* models further, we verified haploinsufficiency in our crispant larvae, showing a significant ∼0.37 fold change (FC) in *ythdf2* expression versus scrambled controls through quantitative RT-PCR analysis (p-value < 0.001; Figure S5). To better characterize *ythdf2* neurodevelopmental phenotypes, we next performed sci-RNA-seq [108] of knockout and overexpression models at 3 dpf, profiling 19,141 single cells from mechanically-isolated heads (average of 4,785 cells per group or 1,126 cells per biological replicate; Table S7). Using known marker genes, we identified nine broad clusters and observed widespread localization of *ythdf2* across cell types (Figure 4G, Figure S6A). This agrees with its reported general expression in humans [109] and zebrafish [110,111] (Figure S7), which begins early in development at 3 hours post fertilization [112].

Differential gene expression analysis across all cells (with counts balanced relative to controls, see Methods) revealed 131 DEGs in our *ythdf2* knockout and 33 DEGs in *YTHDF2* mRNA overexpression model versus respective controls (adjusted p-value cutoff = 0.05, log2FCcutoff = 0.1; Figure 4H, Table S8). While our ASD-DM network did not show associations with DEGs in our models (n=1826; Figure 2), we did observe a significant enrichment of high-confidence Fragile X Syndrome (FXS) protein (FMRP) targets amongst our DEGs (3.6% expected vs 7.2% observed enrichment of DEGs considering 842 target genes; Fisher’s exact test BH-adjusted *p*-value = 0.008; Figure 4H). This list includes genes with known functions in neurodevelopment, including ncam1a, cux1a, unc5a, tbc1d9, ncam1a, camta1a, magi2b, syt1a, sv2bc, stxbp1b.,

Recent studies have suggested that FMRP and YTHDF2 compete for binding to m^6^A-methylated RNA targets impacting their stability [70,113]. Overlapping expression of the 15 FMRP-target DEGs through joint density profiles shows strongest expression across brain cells. Further subtyping the brain cluster into 18 cell types (Figure 4I, Figure S6B) highlights forebrain and midbrain neurons. Considering all FMRP-target genes, we observed significantly reduced expression in *YTHDF2* mRNA compared with *ythdf2* knockout larvae considering all cells (*p*-value = 2.4x10^-7^, Figure 4H) and only brain cells (*p*-value = 2.4x10^-7^, Figure S6C). These combined results are consistent with previous studies implicating YTHDF2 as preferentially binding to FMRP target genes resulting in mRNA degradation and global downregulation.

**Figure 4.**
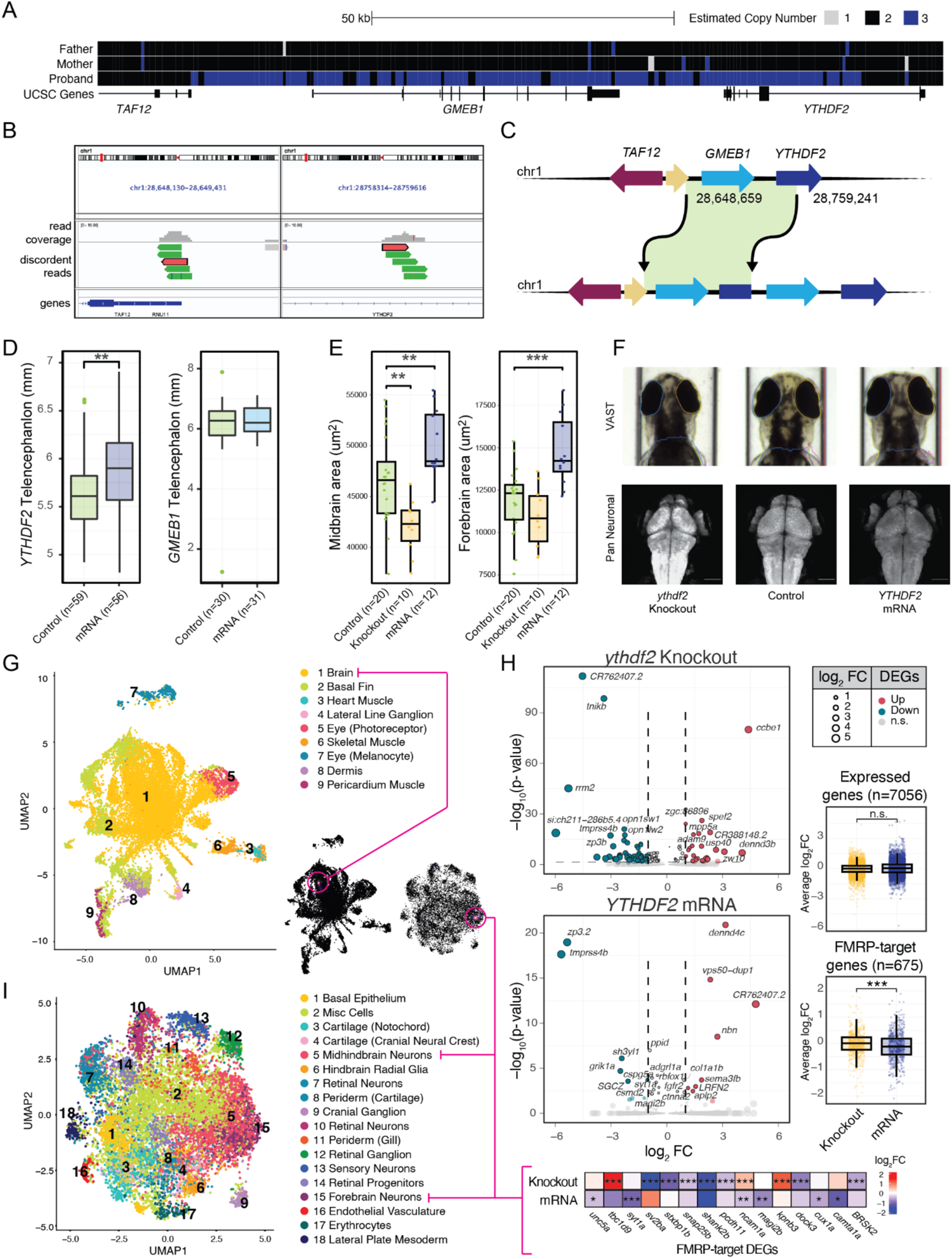
Disrupting *ythdf2* in zebrafish is associated with head and brain size phenotypes. (**A**) Copy-number-estimate plot (QuickMer2) using sequencing data from the APP proband harboring a *de novo* duplication a 109-kb duplication on chromosome 1 compared to their parents harboring two diploid copies. (**B**) IGV plot showing discordant reads in the APP proband supporting a tandem duplication. (**C**) An illustration of the tandem duplication on chromosome 1 in an APP proband encompassing *GMEB1* and all but the last exon of *YTHDF2*. (**D**) Head size of mRNA injected zebrafish, estimated via telencephalon measurement, are significantly increased after *YTHDF2* mRNA microinjection into zebrafish embryos (Wilcoxon T-test p-value = 0.0013), and unchanged in *GMEB1* mRNA injected embryos at 3 dpf (Wilcoxon T-test p-value = 0.8). (**E**) Knockout and mRNA injected zebrafish harboring a pan neuronal marker reveal brain size differences at 3 dpf. Knockout embryos show significantly decreased midbrain volume (Wilcoxon T-test, p-value = 0.003). mRNA injected embryos show both significantly increased midbrain (Wilcoxon T-test, p-value = 0.007) and forebrain (Wilcoxon T-test, p-value = 0.0005). (**F**) Representative control, knockout, and mRNA injected zebrafish images from wildtype embryos imaged with the VAST bioimaging system and HuC embryos harboring a pan neuronal fluorescent marker. Scale bar is 100 μm. (**G**) Hierarchical clustering of 19,141 cells across all conditions into 9 broad cell types based on the expression of gene markers. Joint kernel density estimation was calculated from all 15 FMRP-target DEGs (Nebulosa) highlighting higher expression within a sub-type of brain cells. (**H)** Volcano plots showing DEGs across all cell-types within *ythdf2* knockout (top left) and *YTHDF2* mRNA overexpression (bottom left) conditions relative to controls. DEGs with absolute log_2_FC ≥ 1 and p-adj ≤ 0.05 are colored (upregulated as red, downregulated as blue). Average log_2_FC across all expressed genes shows no significant difference between knockout and overexpression groups (top right), while average log_2_FC across orthologs of all 675 FMRP-target genes expressed in both groups shows a reduction in overexpression relative to knockout. (**I**) Hierarchical sub-clustering of 12,066 brain cells across all conditions into 18 cell types based on the expression of gene markers. Joint kernel density estimation plot highlights higher FMRP-target DEGs expression within forebrain and mid- and hindbrain neurons. Average log_2_FC for all 15 genes across knockout and overexpression with respect to controls is shown. All *p*-values in this figure are represented as: <0.05*, <0.01**, <0.001***.

## DISCUSSION

The autism sub-phenotype ASD-DM, which occurs in approximately 15% of autistic boys, is associated with lower language ability at age three and slower gains in IQ across early childhood resulting in a higher proportion with IQs in the range of intellectual disability by age six [24]. Clues at the underlying etiology of ASD-DM can be found in high-confidence genes such as *CHD8*, a chromatin remodeler important in early brain development [36,114], and *PTEN*, a tumor suppressor gene that functions in cell proliferation [44]. While variants impacting these two genes alone are estimated to contribute to up to 15% of all ASD-M cases [32], a majority of cases remain unsolved. Here, we examined the genomes of 766 ASD-DM and ASD-M trios and quads from the APP and SSC cohorts to identify 153 ASD-DM candidate genes containing *de novo* LGD variants. Ontologies of affected genes largely matched those previously implicated in ASD [115].

When compared with genes implicated in ASD-N and DM-alone, functions related to synapse assembly and long-term synaptic depression, histone methyltransferase activity, cytoskeletal structure stand out in ASD-DM alone (Table S4). To further disentangle mechanisms shared and unique to ASD-DM, we collectively categorized the identified genes from our study and previously published [32] as high-confidence ASD-DM (n=5), ASD-N (n=13), and DM-alone (n=7), as well as those with uncertain disease relevance (n=128) (Table 1). Perhaps unsurprisingly, 16% of high-confidence disease risk genes exhibit recurrence in our cohort, including *CHD8* with three probands affected, while only two genes (*GALNT18* and *LTN1*) in the uncertain “Other” category. These latter genes represent compelling ASD-DM risk candidates, with *GALNT18* (Polypeptide N-Acetylgalactosaminyltransferase 18) functioning in O-linked glycosylation, and *LTN1* (Listerin E3 Ubiquitin Protein Ligase) encoding a RING-finger protein and E3 ubiquitin ligase [36,116–120].

**Table 1.**
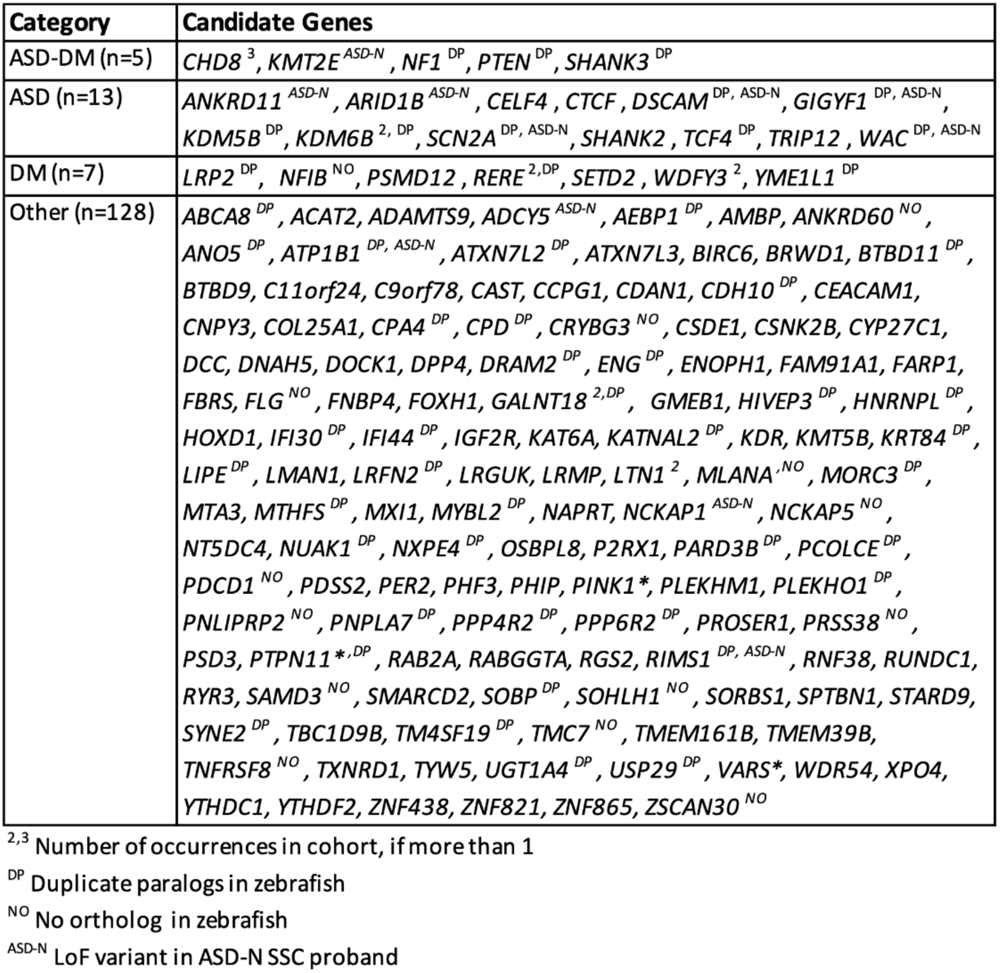
Candidate ASD-DM genes.

As 43% of *de novo* LGD variants in probands have been estimated to contribute to an autism diagnosis, we anticipate that many of the genes identified in this study contribute to ASD-DM or autism in general [9]. Nevertheless, only a single variant was discovered for a majority of candidates limiting our ability to narrow in on true causal genes. As a result, we functionally tested seven genes using zebrafish coupled with CRISPR knockout and discovered two resulting in reduced head sizes in 3 dpf larvae: *ryr3* and *ythdf2*. In the case of the *ryr3* knockout, our findings of microcephaly are counter to those observed in the autistic proband with an identified LGD splice-site variant of *RYR3* resulting in DM (chr15:33634585; G>T). Further experimentation delineating the molecular mechanisms of the variant will be necessary in order to determine if reduced head size in knockout zebrafish larvae might be due to splice alterations of *RYR3* leading to gain-of-function in the patient, or intra-species differences in ortholog functions between humans and zebrafish.

To appropriately model the identified patient *YTHDF2* duplication, for which we hypothesize gene gain-of-function effects, we overexpressed human *YTHDF2* in zebrafish, recapitulating both increased head and brain sizes (Figure 4). To our knowledge, *YTHDF2* gain-of-function has not previously been characterized *in vivo*. Alternatively, published knockout models of the gene in mice [121] and zebrafish [95], using TALENs and morpholinos that target maternal *ythdf2* transcripts, exhibit severe phenotypes and large rates of embryonic death. Generally, the gene exhibits considerable functional constraint between species (e.g., 95% homology with mouse and 72% with zebrafish) and across hundreds of thousands of sequenced humans, with a significant depletion of LGD SNVs discovered to date (gnomAD pLI score of 1, LOUEF score 0.132) [81]. Further, assessment of individuals from the 1000 Genomes Project (n=2,504), SSC families (n=9,068), and gnomAD (n=464,297) did not identify any CNVs impacting *YTHDF2*. Combined, these results highlight the rarity of the 109-kbp duplication impacting *YTHDF2*, identified in a single ASD-DM proband, and suggests that variants impacting this gene could plausibly lead to disease pathogenicity and/or lethality.

In addition to its high conservation, *YTHDF2* is a feasible contributor to ASD-DM based on its previously-implicated functions in neurodevelopment [121,122]. The encoded protein exists in the cytoplasm where it designates m^6^A-labeled RNA for degradation [123] through the recruitment of protein complexes that deadenylate and de-cap mRNA [123,124]. It has over 3,000 target transcripts, including some previously associated with ASD such as *CREBBP* [94]. Studies using induced pluripotent stem cells show that the gene is required for neuronal fate determination, with *YTHDF2* knockdown leading to delayed mitotic entry [125,126] and inhibited pluripotency [127]. Conditional knockout mouse models of *Ythdf2* exhibit decreased cortical thickness as a result of reduced neurogenesis in early development [121]. Together, these results suggest that *YTHDF2* duplication leads to delayed neuronal fate determination, resulting in an overabundance of neuronal progenitor stem cells followed by increased neurogenesis.

While functions of *YTHDF2* in cell cycle and proliferation mechanistically associate it with brain-size phenotypes, evidence of its interactions with mRNA-binding FMRP provide plausible connections with ASD. FXS, caused by loss of FMRP function, represents the most common single-gene cause of ASD, accounting for 2–6% of diagnosed cases [128]. Interestingly, FXS has a similar increased prevalence of macrocephaly as in ASD [129,130]. Several studies have shown FMRP preferentially binds modified RNAs through recognition of m^6^A consensus motifs resulting in protection of transcripts from YTHDF2-mediated degradation [131–133], possibly through direct interactions of the two proteins. This is evident in mouse neuroblastoma cells, where the loss of Fmrp is associated with reduced m^6^A-modified transcripts, while knockdown of *Ythdf2* leads to increased stability and longer half lives of modified RNAs [131].

In our transcriptomic analysis of *YTHDF*2 zebrafish models, we found significant enrichment of FMRP-target DEGs when considering both knockout and mRNA-overexpression larvae, with several genes exhibiting opposing effects in knockout versus overexpression (Figure 4H). One example is *ncam1* (Neural adhesion molecule 1), where we observed significant upregulation in *ythdf2*-knockout and downregulation in *YTHDF2*-mRNA larvae; this is in line with several studies in humans connecting depressed *NCAM1* expression with ASD [134–137]. Further, we show that overexpressing *YTHDF2* in zebrafish is associated with decreased expression of FMRP-target genes across all cells and in the brain (Figure S6), likely due to increased degradation of m^6^A-modified mRNAs. Based on these collective findings, we propose a model in which *YTHDF2* loss-of-function results in microcephaly and possibly mortality, by increasing stability of m^6^A-labeled RNAs resulting in extended cell-cycle progression and a reduction in neurogenesis (Figure 5). Alternatively, *YTHDF2* gain-of-function results in increased degradation of m^6^A-modified transcripts, possibly contributing to megalencephaly through increased neurogenesis and ASD through haploinsufficiency of FMRP-target genes [138].

**Figure 5.**
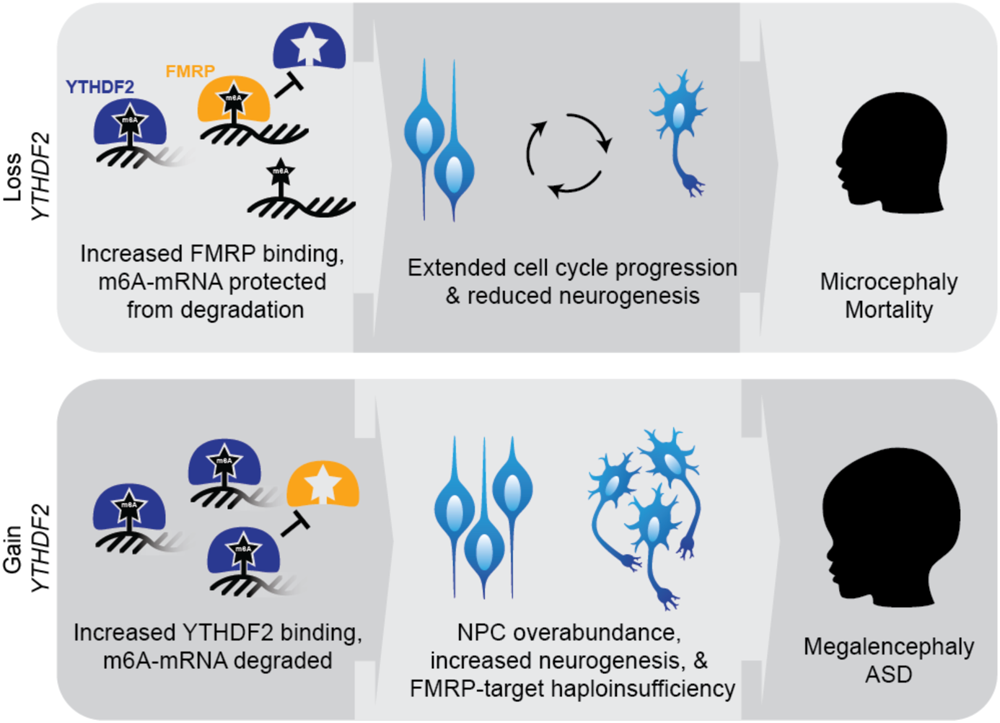
A potential role for *YTHDF2* in ASD-DM. Proposed model of *YTHDF2* loss- or gain-of-function phenotypes, with respect to FXS protein FMRP. We hypothesize *YTHDF2* loss-of-function would lead to microcephaly due to increased FMRP binding and lack of m6-mRNA degradation, extended cell cycle progression, and reduced neurogenesis. As the gene is highly conserved and knockout models are embryonic lethal, likely loss-of-function mutations in humans lead to disease pathogenicity or are incompatible with life. Inversely, *YTHDF2* duplication would lead to megalencephaly following increased m^6^A-mRNA degradation as YTHDF2 outcompetes FMRP, neural progenitor cell (NPC) overabundance, and increased neurogenesis.

Expanding beyond *YTHDF2*, we identified an additional *de novo* LGD variant in an SSC ASD-M proband impacting another YTH-domain-containing m^6^A-RNA reader, *YTHDC1*. The encoded protein promotes recruitment of splicing factors and facilitates nuclear export of modified transcripts [139]. Where gain of *YTHDF2* may lead to a decrease in overall m^6^A-mRNA by promoting transcript degradation, conversely the loss of *YTHDC1* would likely result in a depletion of spliced and cytoplasmic transcripts [140]. Interestingly, FMRP also facilitates nuclear export of m^6^A-labeled RNAs prevalent during neural differentiation [132,141,142]. While m^6^A-RNA regulation genes were not significantly enriched in our identified ASD-DM candidate genes, none were observed in SSC ASD-N or in TD siblings [143], suggesting that additional genes within this pathway may contribute to ASD-DM. Indeed, FMRP has been shown to repress translation of m^6^A RNAs, through competition for binding and inhibition of m^6^A reader *YTHDF1* [144]. These ties between m^6^A mRNA readers and FMRP suggest a fine balance of select transcripts, in which up- or down-regulation may impact early neurodevelopment and autistic phenotypes. Further, the m^6^A-labeled mRNA flavivirus ZIKA is associated with severe congenital microcephaly, with *YTHDF2* found to bind and destabilize viral RNA [145,146]. These antiviral functions are controlled in part by the *METTL3* methyltransferase, which labels viral RNA for degradation, and whose knockout models are also associated with a reduced brain size in mice [147]. Together the m^6^A-mRNA pathway—including YTH-domain proteins and m^6^A de/methyltransferases — represents a compelling future area of study in regard to ASD and brain-size phenotypes.

Though new insights were achieved from our study, we would like to highlight some limitations. Due to the dearth of MRI evidence, not all SSC ASD-M probands included in this study will meet the criteria for ASD-DM. We do expect the overlap to be significant, as macrocephaly has previously been found to be highly correlated with megalencephaly, with an increased correlation in young children [148]. This is supported by the majority of APP ASD-DM probands also meeting the criteria for macrocephaly (82%), Additionally, zebrafish, with a forebrain that most closely resembles the mammalian neocortex [149], may not be a suitable model for all ASD-DM candidate genes. This is highlighted by our microcephaly finding in *ryr3* knockout zebrafish larvae, counter to the APP proband phenotype. Further, our study did not identify any measurable morphometric differences in *chd8* crispants versus controls at 3 dpf (Figure 3), counter to previous studies showing megalencephaly in crispants, morpholino knockdown, and stable KO lines at young larval stages [36,96]. We do note that inconsistencies exist across published *CHD8* KO models, with a recent stable line reported to show decreased brain size at 6 dpf [35]. Nevertheless, characterizing genes implicated in ASD and other neurodevelopmental conditions in zebrafish has been successfully demonstrated across hundreds of genes (reviewed by [150–154]).

Overall, this study represents a significant increase in the number ASD-DM and ASD-M proband genomes analyzed in search of candidate genes. The 153 candidate ASD-DM genes introduced here greatly expand our knowledge of the genetic factors specifically contributing to this severe subphenotype of ASD. With this expanded list of ASD-DM candidate genes, network analysis can now be leveraged to identify additional candidate genes with similar gene functions to known ASD-DM genes. Our study introduces two novel ASD-DM candidate genes connected with head-size phenotypes in a zebrafish model system and 141 novel unvalidated ASD-DM candidate genes (Table 1). Finally, our research establishes a roadmap for the rapid functional characterization of these putative risk genes, demonstrating the methodology needed to validate these candidates going forward.

## Acknowledgements

Thank you to the Autism Phenome Project (APP) research staff, especially Dr. Brianna Heath, and clinical recommendations from Dr. Suma Shankar, as well as the undergraduate students that provide husbandry and care for our zebrafish. We are grateful to Dr. Bruce Appel from the University of Colorado for sharing the HuC-eGFP line. A special thank you to the families who have generously shared their genetic data – you are who all of this research is for.

## Data availability statement

Raw sequencing data of patients, including FASTQ and VCF files, can be accessed through the MSSNG access agreement (https://research.mss.ng) and the Simons Simplex Collection through SFARI Base (https://www.sfari.org/resource/sfari-base/). Transcriptomic data from zebrafish mutants is available through NCBI GEO (Accession # pending).

## Funding statement

This study was supported by a pilot grant from the UC Davis MIND Institute Intellectual and Developmental Disabilities Research Center funded by NIH National Institute of Child Health and Human Development (P50HD103526), the NIH National Institute of Neurological Disorder and Stroke (R21NS128811), and the NIH Office of the Director and National Institute of Mental Health (DP2MH119424) to MYD. SSN is supported by the NIMH Autism Research Training Program T32 (MH073124) through the UC Davis MIND Institute; NAFM is supported by the NIGMS as a UC Davis Postbaccalaureate Research Education Program fellow (R25GM116690); GNL is supported by an NINDS Research Supplement to Promote Diversity in Health-Related Research (R21NS128811-01A1W1); NKH is supported by an NIGMS UC Davis eMCDB T32 (T32GM153586).

## Conflict of interest disclosure

Authors have no conflicts of interest to report.

## Clinical trials

This study involves no clinical trials. In accordance with the ICMJE and to be considered for review in Autism Research, the journal requires that clinical trials are prospectively registered in a publicly accessible database and clinical trial registration numbers are included in all papers that report their results. A clinical trial is defined as any research study that prospectively assigns human participants or groups to one or more interventions to evaluate the effects of those interventions on health-related biomedical or behavioral outcomes. The registry must be open to all registrants and managed by a not-for-profit group, and must have a mechanism to guarantee accuracy and validity of the information submitted. The registry must adhere to the ICMJE mandates described in the table found on their website. For more information regarding what Autism Research considers to be a clinical trial, please visit the Clinical Trial Registration section of our author guidelines.

## Notes

### Competing Interest Statement

The authors have declared no competing interest.

### Summary of Updates

Additional experiments were added characterizing single-cell transcriptomes of YTHDF2 zebrafish models. As a result, we found connections with FMRP-target genes in our knockout and overexpression models, providing insight into etiology of ASD-DM in our proband.

