## Supplementary Figures for "m^6^A-mRNA reader *YTHDF2* identified as a potential risk gene in autism with disproportionate megalencephaly"

#### SUPPLEMENTARY TABLES

**Table S1.** CRISPR zebrafish knockout gRNA and primer sequences.
**Table S2.** ASD-DM and ASD-M probands from the SSC and APP cohorts. DM, disproportionate macrocephaly; RM, relative macrocephaly; SO, somatic overgrowth.
**Table S3.** Loss-of-function *de novo* variants from ASD-DM and ASD-M probands.
**Table S4.** Enriched gene ontologies from network analysis of ASD-DM candidate genes v a human background from DAVID.
**Table S5.** Network analysis seed ASD-DM candidate genes and top 10 interactors.
**Table S6.** Enriched gene ontologies from network analysis of ASD-DM candidate genes and top 10 interactors.
**Table S7.** Technical statistics for scRNA-seq replicates.
**Table S8.** Differential gene expression results from scRNA-seq and FMRP-gene assignment.

### SUPPLEMENTARY FIGURES

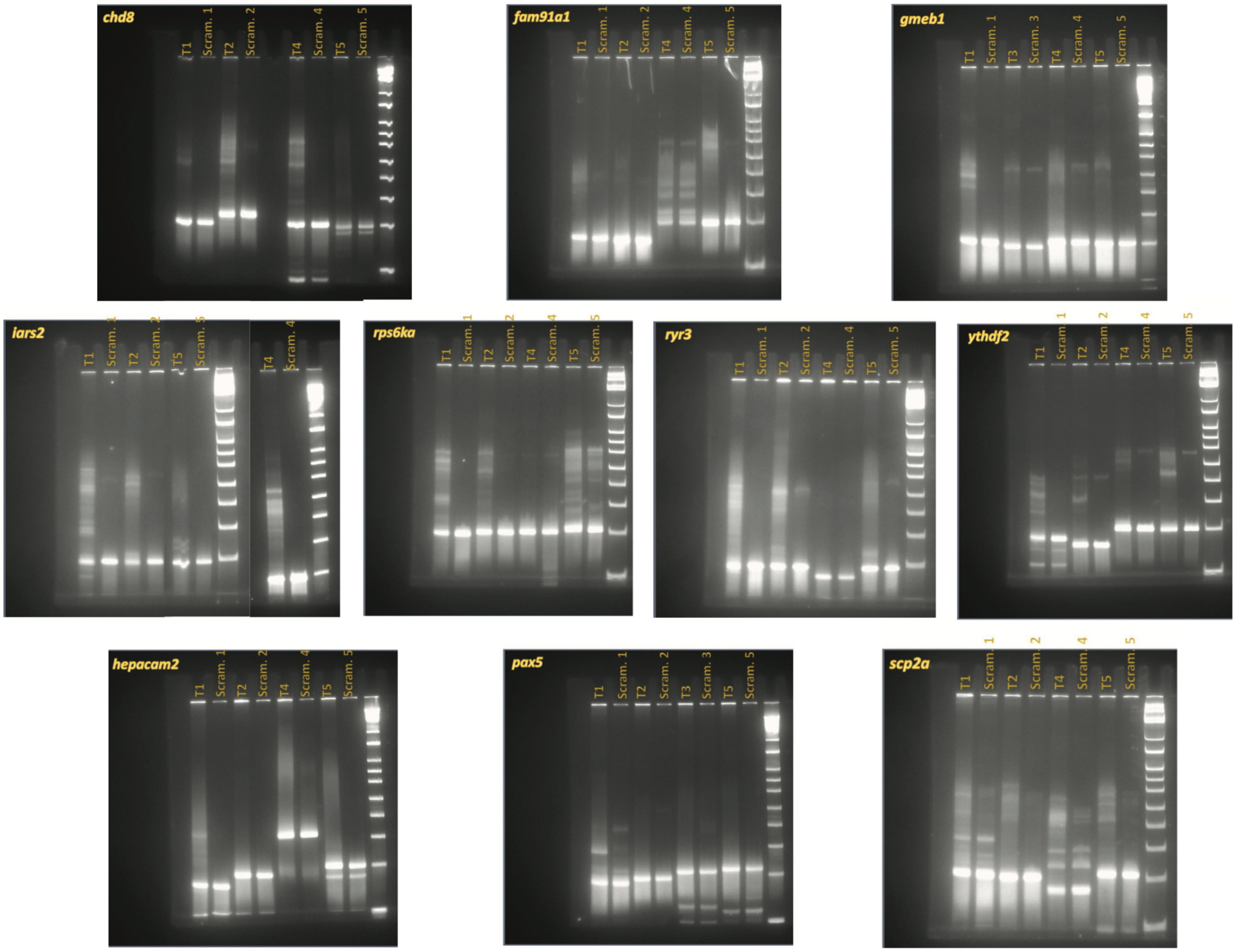


**Figure S1. Mutational efficiencies of CRISPR targets in zebrafish embryos.** Polyacrylamide gels showing the efficiency of each of the four CRISPR gRNAs chosen for the 10 genes tested in zebrafish.

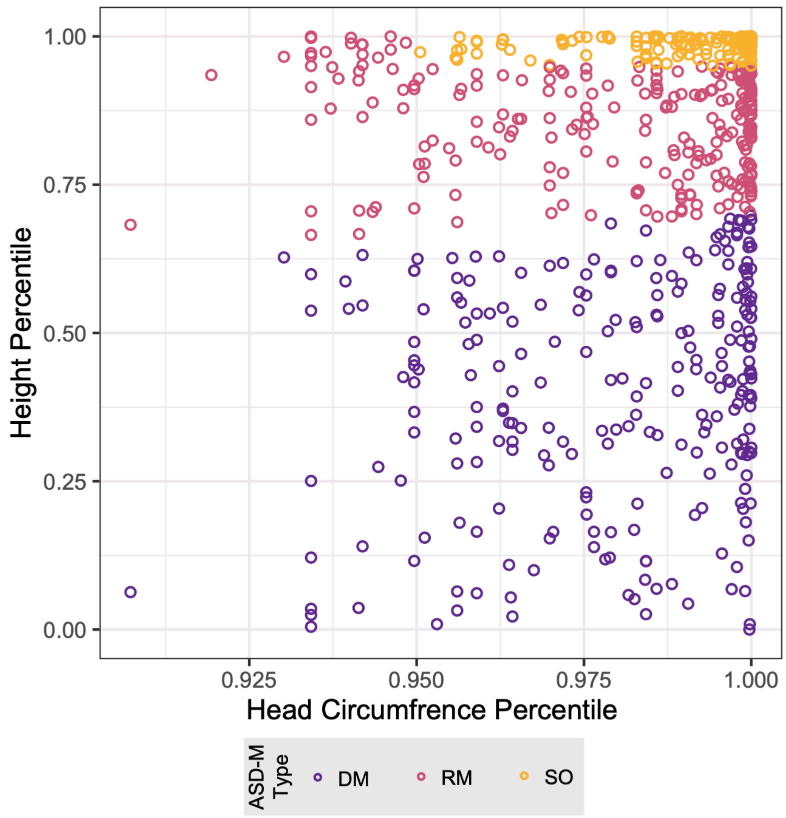


**Figure S2. Head circumference v. height percentile of ASD-M SSC probands.**

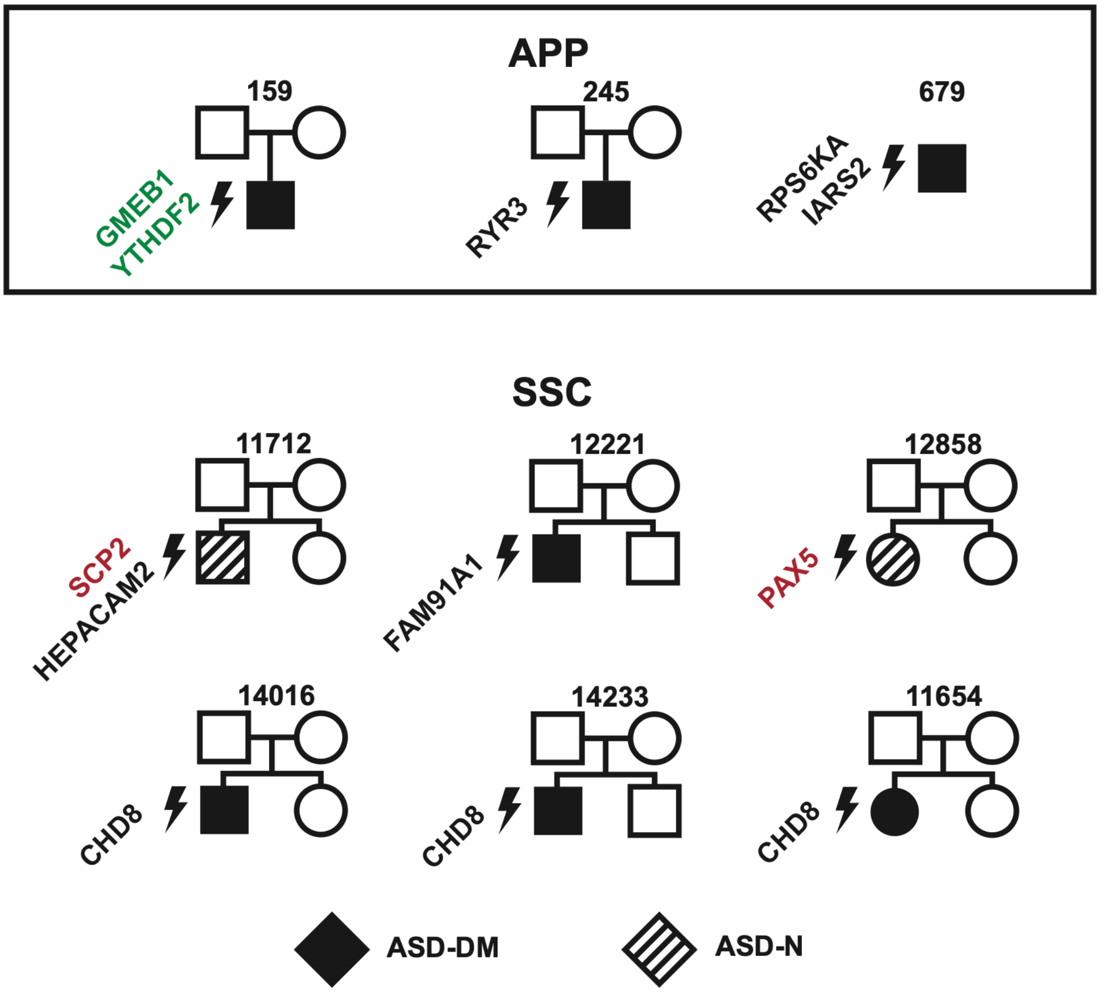


**Figure S3. ASD-DM and ASD-M families with *de novo* variants.** Families were from the Autism Phenome Project (APP) and the Simons Simplex Cohort (SSC) with impacted genes that were screened using zebrafish in this study. Green font signifies a duplication and red font signifies a deletion.

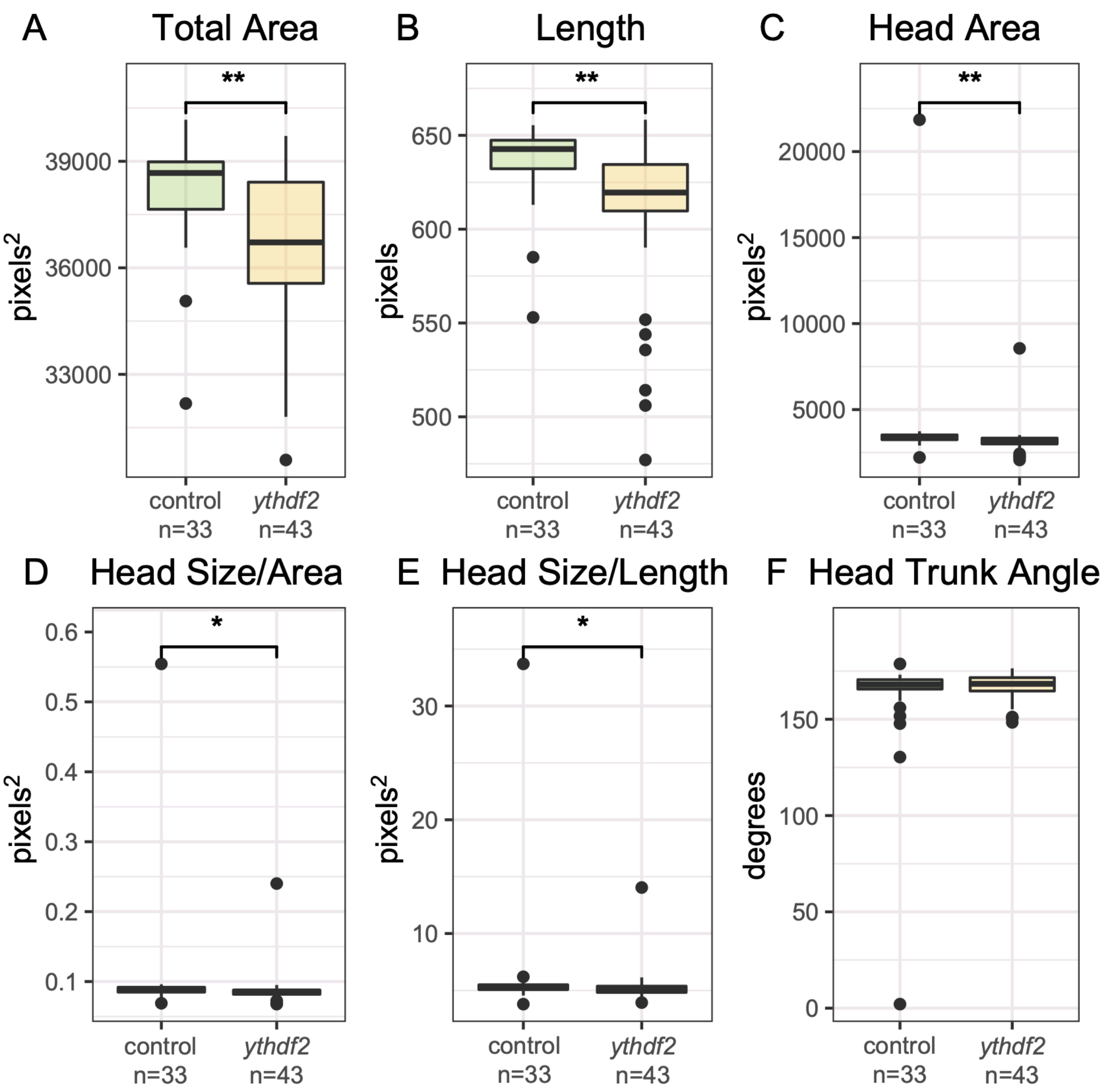


**Figure S4. Lateral VAST measurements of *ythdf2* knockout embryos at 3 dpf.** Measurements of total area (**A**), length (**B**), and head area (**C**) were significantly reduced (Wilcoxon T-test $p$-value < 0.00041), with a significantly reduced head size maintained after normalization by area (**D**) (Wilcoxon T-test $p$-value = 0.032) and length (**E**) (Wilcoxon T-test $p$-value = 0.044). No significant change noted in head trunk angle (**F**) (Wilcoxon T-test $p$-value = 0.56). These results are consistent with overall smaller embryos and microcephaly, but not developmental delay.

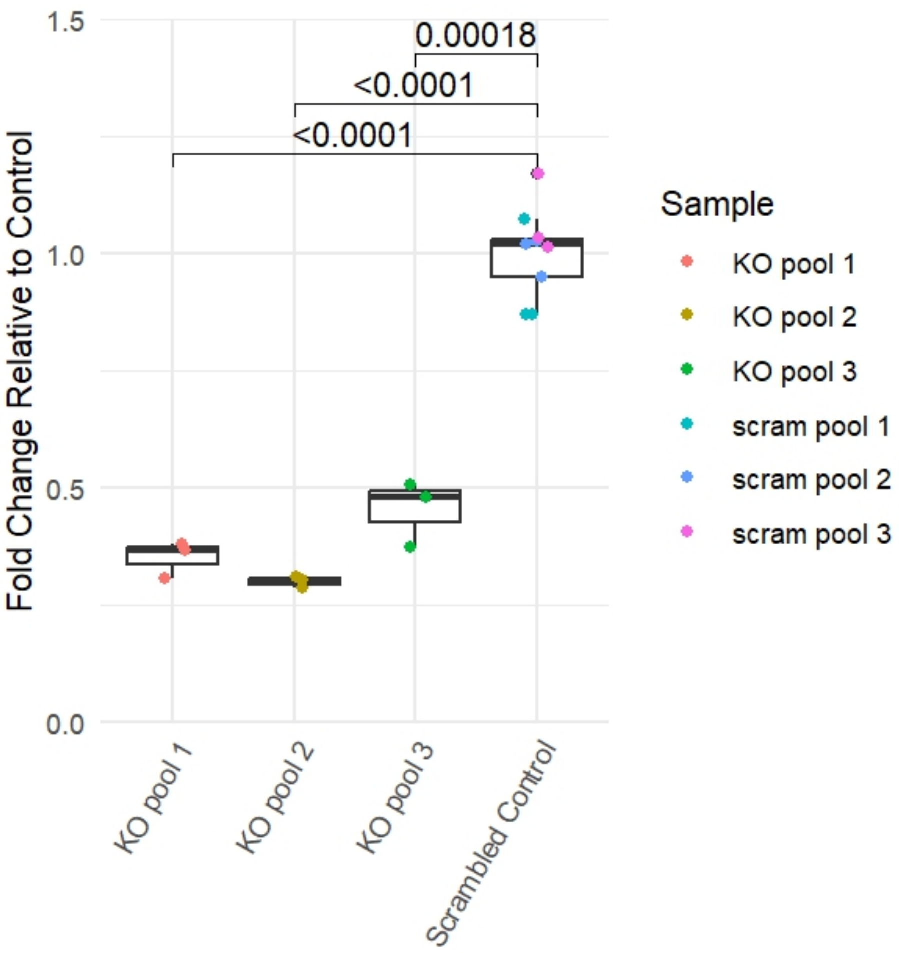


**Figure S5. Gene expression analysis of *ythdf2* in crispant mutants.** RT-qPCR results for  across *ythdf2* knockout (KO) and scrambled control biological replicates. All samples demonstrate consistent *act1b* expression, while the pooled *ythdf2* KO samples demonstrate lower *ythdf2* expression than the scrambled control. *p*-values were calculated using a Student's T-test.

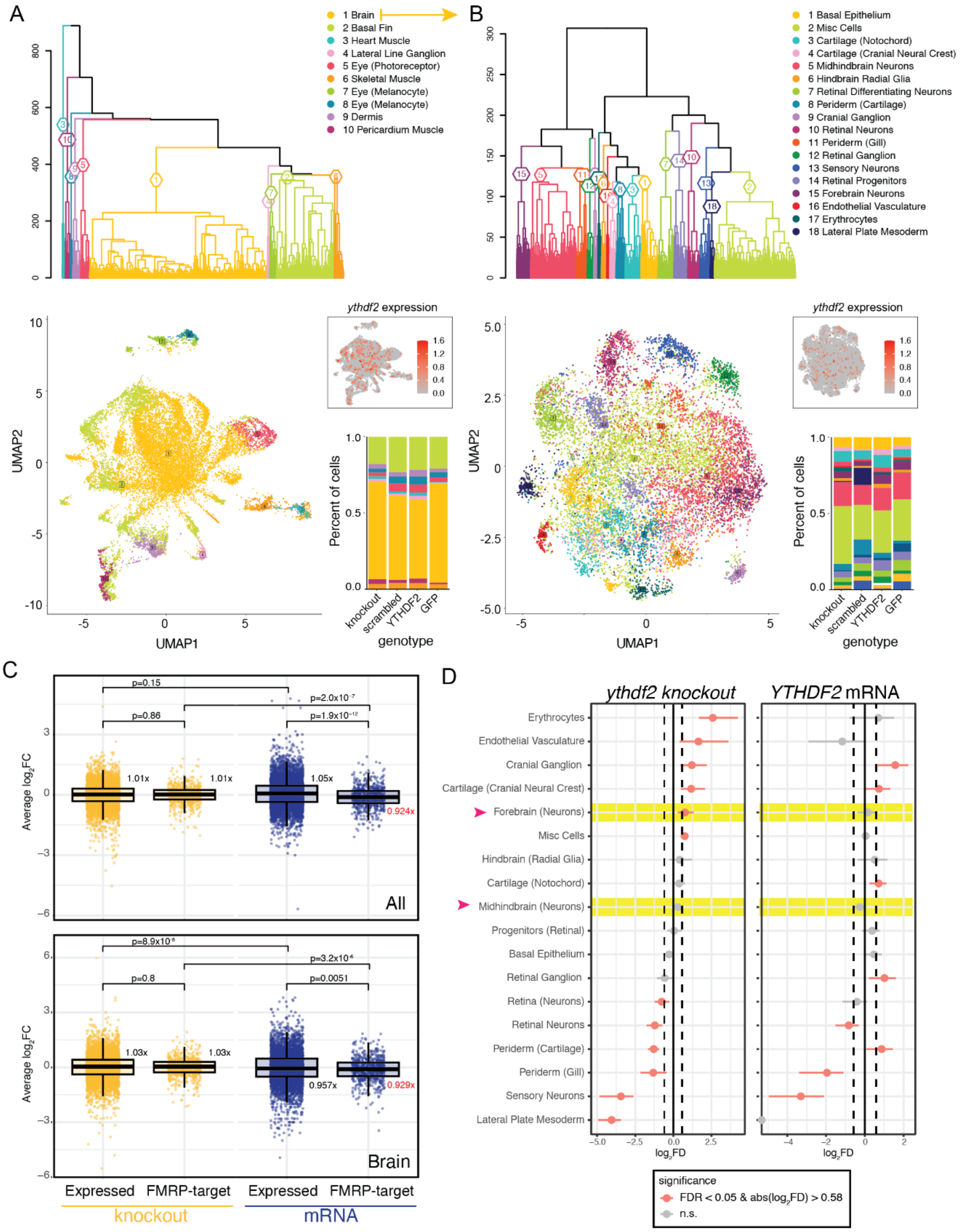


**Figure S6. Single-cell transcriptomic analysis of *YTHDF2* zebrafish models.** Dendrograms were created to cluster cells with similar expression profiles and visualized via a UMAP plot, with colors indicating assigned cluster IDs for (**A**) all cells and (**B**) brain cells assigned in A. The proportion of cells assigned to each cluster per model is a percentage of total cells indicated as a barplot. Cells with *ythdf2* transcripts are colored red based on a continuous scale of natural log normalized expression [1]. (**C**) The average $\log_2$FC per expressed gene (0.01% of cells) was plotted across all cells and brain cells (see Methods) for all and FMRP-target genes for the *ythdf2* knockout and *YTHDF2* mRNA zebrafish models. Comparisons were made using T-tests either paired (between models) or unpaired (within models). Median fold changes versus respective controls (scrambled for knockout and GFP for mRNA) are indicated next to plots. (**D**) Abundances of cells per brain subcluster were plotted as $\log_2$ fold difference (FD) with error bars representing 95% confidence intervals and an FDR threshold of 0.05 (scProportionTest).

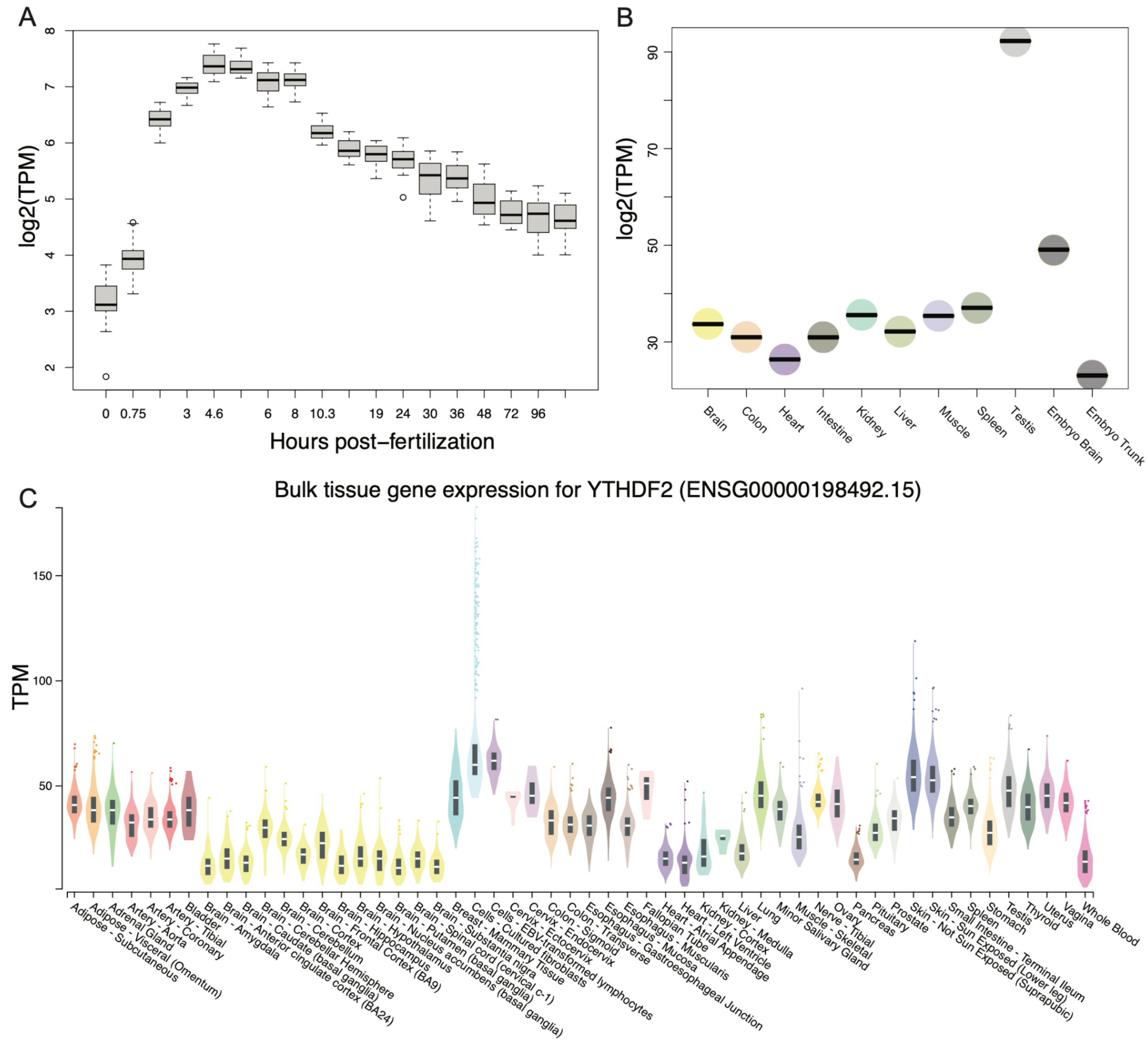


**Figure S7. Expression of *YTHDF2* in zebrafish and humans.** (**A**) Expression of ythdf2 showing increased transcript per million (TPM) over early developmental time, peaking at ~5 hours post fertilization [2]. (**B**) Bulk tissue expression of *ythdf2* in zebrafish showing broad expression, highest in adult testis and embryo brain [3]. (**C**) Bulk tissue gene expression of *YTHDF2* from the Genotype-Tissue Expression project (GTEx) showing high broad expression in humans [4].
